# Mechanisms linking Adverse Childhood Experiences to adolescent mental health: a participatory arts-based study of adolescents’ accounts

**DOI:** 10.64898/2026.07.30.26359340

**Authors:** Siobhan Hugh-Jones, Chloe Farahar, Luke Allder, Annette Foster, Emma Williams, Kamaldeep Bhui, Nicola Shaughnessy

**Affiliations:** School of Psychology, University of Leeds, UK; School of Arts and Architecture, University of Kent; School of Psychology, University of Surrey; Department of Psychiatry, University of Oxford

**Keywords:** adverse childhood experiences, adolescent mental health, arts-based participatory research, trauma-informed research, psychological safety

## Abstract

Research on adverse childhood experiences (ACEs) has largely relied on retrospective and predominantly adult-focused models that conceptualize mental health difficulties as outcomes of past adversities operating through linear causal pathways. Less is known about how adolescents themselves understand the mechanisms linking adversity and mental health. This study explored young people’s lived experiences of these mechanisms using participatory arts-based methods. Sixty-two young people aged 10-24 years from diverse and often underrepresented backgrounds across England participated in trauma-informed creative workshops. Workshops incorporated multiple artistic modalities, including visual arts, animation, drama, dance, music, film, and creative writing, generating experiential and conversational data. Data were analysed using Framework Analysis within a critical realist approach. Young people did not primarily describe their mental health through narratives linking past adverse events to current outcomes. Instead, they emphasized present-day relational, environmental, and institutional conditions as the most salient influences on wellbeing. Two interconnected pathways were identified: system failures and seeking restoration. System failures referred to ongoing experiences of invalidation, bullying, sensory overwhelm, masking of identity, and unresponsive educational or mental health systems that generated feelings of unsafety. Seeking restoration encompassed actively pursued experiences of belonging, community, validation, sensory regulation, nature connection, creative expression, trust, and authenticity that supported wellbeing. Across pathways, felt (un)safety emerged as the central organizing mechanism through which experiences affected mental health. Findings suggest that adolescents explain their mental health less in terms of historical adversity and more through current experiences of safety, recognition, and belonging. Trauma-informed research and practice may therefore benefit from complementing questions about past adversity with greater attention to what is happening in young people’s lives now and the conditions that support recovery and flourishing.

## Adverse Childhood Experiences and Adolescent Mental Health

Prevalence rates of mental ill-health during adolescence are of concern globally (McGorry et al., 2024). Although adolescence is a developmental period during which many mental health difficulties first emerge, access to timely, effective, and non-stigmatising support remains limited for young people in most countries (Ghafari et al., 2022). In response, national policy agendas increasingly emphasise preventative public mental health approaches, which require identification of modifiable intervention targets in youth populations (Papola et al., 2024). These targets are typically among the social, relational, and environmental determinants of adolescent mental health rather than in individual pathology. Adverse childhood experiences (ACEs) and their sequelae are widely recognised as pervasive, significant, and in principle avoidable, social determinants of adolescent mental health and therefore a critical target for prevention approaches (Danese, 2020).

ACEs are very challenging and potentially traumatic events occurring before the age of 18 years that can have lasting effects on mental, physical, social and economic wellbeing across the life course (Obse et al., 2026; Sahle et al., 2022). The first identified ACEs were abuse (physical, sexual, or emotional), neglect (physical or emotional), and household difficulties such as parental separation, substance misuse, mental illness, or incarceration (Felliti et al., 1998). Subsequent studies with more diverse populations identified other ACEs that are comparable, if not stronger, predictors of harm and lifelong impacts. These are bullying, community violence (including racism and discrimination), neighbourhood unsafety, disability, poverty, and involvement in foster care or child protection systems (Asmussen et al., 2021; Cronholm et al., 2015). Many, though not all ACEs, will mean that a child has experienced trauma. Taken from the ancient Greek term for bodily wound, the psychological definition of trauma continues to evolve (e.g. DSM-V-TR, APA 2022; Gradus & Galea, 2022) but can broadly be defined as an experience or event that overwhelms an individual’s ability to cope (Zoromba et al., 2024).

Although estimates vary by population and measurement approach, evidence shows that many people experience ACEs. Based on global data using the originally identified list of ACEs, approximately 22% report one ACE, 13% two ACEs, 8% three ACEs, and 15% four or more ACEs (Madigan et al., 2025). Comparable and sometimes higher rates have been reported in UK samples (the site of our study) (Bellis et al., 2015; Houtepen et al., 2020). Experiencing ACEs is robustly associated with adverse outcomes across the life course, and intersectionalities, with risk increasing with each additional ACE (Boullier & Blair, 2018; Havers et al., 2024; Hughes et al., 2017). Impacts on mental health can be profound and enduring, often due to the impact of trauma (e.g. intrusive thoughts, nightmares, flashbacks, emotional numbing, hyperarousal, relationship difficulties; Straussner & Calnan, 2014). Experiencing childhood adversity and trauma is also associated with later anxiety, depression, substance use, post-traumatic stress disorder, self-harm and suicidality (Bellis et al., 2014; Lewis et al., 2019). Some population-attributable estimates suggest that ACEs account for roughly 30% of anxiety and 40% of depression cases in North America, and over a quarter of anxiety and depression cases in Europe (Bellis et al., 2015).

The most effective strategy for preventing poor adolescent mental health outcomes associated with ACEs is the primary prevention of adversity and trauma itself (Brennan et al., 2021). However, given the structural and social conditions that make ACEs difficult to entirely eliminate, a critical additional approach is to identify the risk and resilience mechanisms through which ACEs influence developmental trajectories and outcomes. Such knowledge would enable the targeting of timely, developmentally informed interventions at key points where mental health risk may be amplified or mitigated (Baker et al., 2021).

### Mechanisms Linking ACEs to Mental Health Outcomes

Considerable mechanistic understanding of how ACEs shape later adult mental health has been established. This body of work highlights the role of complex, interacting pathways involving stress-regulation systems, emotion regulation, threat sensitivity, and relational functioning (Danese, 2020). However, much of the mechanisms and outcomes evidence is adult-centric and does not tend to consider adolescent data or their lived experiences. This is partly due to methodological challenges and biases. Many studies with adolescents use standardised measures of ACE that were originally developed for adult populations. Such measures were designed to capture retrospective, cumulative exposure across an entire childhood, assuming stable memory, narrative coherence and subsequent adult meaning-making. When applied to adolescents, whose ACEs are more recent or potentially ongoing, and during a time of considerable personal and social change, these measures may misrepresent exposure, underestimate prevalence, or fail to capture contemporary and developmentally salient adversities (e.g. online harm, global conflict) (Lam et al., 2024).

In addition, the high reliance on parental reports of adversity on behalf of a young person is also problematic, given divergence in accounts and the importance of knowing the individual’s subjective experience and meaning making of the adversity (Baldwin et al., 2019). Furthermore, whilst there has been a predominance of studies on the associations between ACE exposure and outcomes using checklist-based, event-focused reporting, there are relatively few which examine subjective experience of ACEs and what this can tell us about the mechanisms leading to differential adolescent mental health outcomes. Finally, whilst some work has tried to identify resilience factors following childhood adversity, these are often driven by theoretical enquiry rather than by lived experience (e.g. Fritz et al., 2018; Schaefer et al., 2023). Compared to adults, we know relatively little about how adolescents interpret, negotiate, and adapt after or during ongoing adversity and trauma, and why some develop mental health difficulties while others do not (Danese, 2020). We are thus lacking the contextualized, lived experience knowledge that might effectively inform real-world interventions to mitigate the long-term mental health impacts of ACEs.

### The Need for Lived Experience data from Diverse Young People

Lived experience as a form of knowledge refers to insight generated from first person accounts of direct, embodied and situated experiences of everyday phenomena and the meanings they ascribe to these experiences within specific social, cultural and historical contexts (Casey, 2023). Typically gathered in the form of qualitative data, lived experience constitutes a legitimate and distinct epistemic resource that complements other forms of knowledge, acknowledging that we require different perspectives to draw out or illuminate what is invisible to others (Casey, 2023). It is one thing to know facts about trauma and its sequelae, but something else entirely to know what it is like to be traumatised and what would have helped as you tried to navigate life after it. Sonuga-Barke (2024a) argues that lived experience data, and broader openness to methodological and knowledge pluralism, is now essential if we hope to advance child psychology and psychiatry and achieve translational impact for mental health. There are now many illustrative examples demonstrating how lived experience data can challenge established assumptions and drive care and interventions about, for example, ADHD (Batura, 2024), psychosis (Fusar-Poli et al., 2022) and eating disorders (Gupta, 2026). Similarly, generating lived experience data from young people with ACEs may offer fresh insight into mechanisms influencing whether adversity is or is not translated into mental health risk during adolescence. Lived experience research may illuminate how young people actively and agentically navigate their lives following adversity, what subsequent experiences, events and conditions matter to them, and where support and prevention opportunities are evident to them and could be harnessed by public mental health approaches (Kim & Royale, 2025; Sonuga-Barke et al., 2024b).

A number of priorities should inform lived experience research on ACEs, trauma and youth mental health (Lane et al., 2025). Given that mental ill-health is unequally distributed and disproportionately affects young people experiencing marginalisation (e.g., related to ethnicity, gender and sexual identity, neurodivergence, and socioeconomic or educational position, Leyreya et al., 2024; Murthy, 2022), research must reach and engage these populations to ensure that the evidence base informing policy and practice reflects the realities of their experiences and lives (Perowne et al., 2024). Addressing patterns of under-representation requires the careful and intentional design of research that is meaningful, inclusive and accessible (logistically, developmentally, relationally and psychologically) to diverse young people with differing identities and from different communities.

Research in this space also needs to recognise traditional lived experience methods, such as interviews, may be challenging, insufficient or excluding for some people. This is because adversity and trauma can create challenges in memory and narrative coherence, attention and emotional expression, and there may be distrust of what may seem like extractive approaches by people in power (Rahapsari & Levita, 2025). Lived experience in this field must therefore be designed in line with trauma-informed research principles (Alessi et al., 2023). This means designing research practices, and cultivating researcher-participant relationships, that seek to avoid unwittingly retraumatising participants or replicating the misuse of power that they may have experienced in their lives via people, communities and / or institutions. It also means carefully supporting the recounting but not reliving of ACEs. Methods are needed which enable young people to express lived experience in ways that are safe for them, are responsive to their capacities and preferences, that resist epistemic injustice through silencing or simplification, and that do not presume coherence, narrativity or verbal fluency as preconditions for meaningful participation (Alessi et al., 2023). A promising way to deliver such research is through participatory arts-based approaches. We set out the rationale for our use of these methods in a separate paper (Pavarini et al., 2021) and we specify below how these were rendered trauma-informed in our study.

### Purpose of the Present Study

The aim of the present study was to examine mechanisms linking ACEs to adolescent mental health through creative ways of eliciting and studying adolescents’ lived experience and meaning-making about the adversity and trauma they endured. Out study was nested within the Attune Project, a multisite study in England (2021-2026) funded by the UK Research & Innovation research council (Bhui et al., 2026). Project Attune’s overarching aim was to discover novel risk and resilience mechanistic pathways linking ACEs to adolescent mental health outcomes, and to identify and co-create targets and resources for early intervention and public health prevention approaches. As a first trauma-informed research principle (Alessi et al., 2023) is to learn with and about the community being researched. the Attune project established a national Young People’s Advisory Group, and multiple regional youth advisory groups, for project duration. Populated by young people with ACEs, they helped us to think about the likely needs and concerns of our adolescent participants, how to design research practices that would promote safety, trust and equitable inclusion (see Batool et al., 2026 for our paper co-written with young people about their experience on our advisory groups). This paper presents the findings of the first study in Project Attune which drew upon trauma-informed arts-based methods (ABMs) to understand adolescents’ lived experiences of ACEs.

In research, ABMs refer to the use of creative practices (e.g., collage, drama, drawing, film and media, movement, music, photography and storytelling). These can be embedded with trauma-informed principles that prioritise transparency, careful pacing, trust-building, choice, relational and empowerment (Pavarini et al., 2021). When delivered in this way, ABMs can acknowledge the psychological, relational, and bodily effects of trauma and create opportunities for individuals to safely express lived experience (with or without accompanying interview approaches) without high risk of re-traumatisation. Evidence shows that ABMs can yield insights not accessible through conventional methods, particularly when working with trauma affected young people (Coholoic, 2011). This may be because ABMs can offer emotional distance, support communication of experiences that are difficult to verbalise or place in a narrative format, and because they reduce cognitive or linguistic barriers, supporting inclusion of groups often excluded from traditional research (e.g., refugees) (Fraser and Galinsky, 2010). Although our use of ABMs has similarities with arts-based phenomenological research (Gupta & Zieske, 2024) given we were interested in the essence of experience, we prioritised a broader account of contextualised lived experience. This was because of our ambitions to generate actionable knowledge for public mental health approaches.

Thus, using ABMs, our aim was to generate new insights into adolescents’ lived experience of mechanisms they felt linked their ACEs to their current mental health. Six research questions underpinned our study, although this paper reports the findings of one in detail. For context, our research questions were: (1) How do young people define what an ACE; (2) What characterises their experience of them?; (3) How do young people affected by ACEs define and explain their mental health?; (4) What are the mechanisms they believe are driving the ways ACEs come to affect their mental health?; (5) What or where do young people believe are the prevention and intervention opportunities to mitigate the impacts of ACEs on adolescent mental health?; and (6) what is the role of creative practices in research on ACEs and in the lives of these young people? The present paper reports the findings focused on research question (4).

We situate our work within a critical realist philosophical framework, which aligns with our use of Framework Analysis as an analytic method (Mercier et al., 2023). Critical realism is characterised by an ontologically realist and epistemologically relativist stance and distinguishes between an independently existing physical and social world and the partial ways in which it is known. It stratifies reality into three domains: the real, comprising underlying structures and generative mechanisms (e.g. overstretched mental health services, racism, neurobiological stress responses); the actual, referring to events and processes that occur when such mechanisms are activated (e.g. feeling unsupported, experiences of trauma or neglect); and the empirical (i.e. what aspects of the actual is observed or reported) (Groff, 2024; Longhofer & Floersch, 2012).

In our study, young people’s narratives were understood as expressions of lived reality situated primarily at the empirical level, accessed through our ABMs. These accounts were treated as highly valid and indispensable sources of knowledge about events and processes operating at the actual level, and as critical clues to the underlying mechanisms constituting the real. At the same time, consistent with a critical realist epistemology, we recognise that lived experience accounts are partial accounts of the real, necessarily mediated by perception, context and meaning-making. Accordingly, our analytic approach involved theorising beyond experience to develop explanatory accounts of why particular outcomes occur and how social, relational and psychological mechanisms operate in shaping youth mental health following adversity.

## Method

We chose participatory ABMs, embedded with trauma-informed practice and relational ethics (Pavarini et al., 2021). This was delivered as a series of facilitated, recorded practical workshops, tailored to different groups of young people. The process of producing creative and performing arts outcomes functioned as a relational and symbolic medium through which lived experience of ACEs could be made visible and communicable.

### Research Team

Mirroring the wider Attune project, the present study was delivered by an interdisciplinary team of senior and early career researchers and creative arts practitioners, spanning the arts and humanities, social science (psychology), and medicine (psychiatry). Operationalising our approach to relational ethics (Pavarini et al., 2021), workshops were designed and facilitated by those experienced in trauma-informed, creative practice with vulnerable young people, including those who are neurodivergent and/or have specific communication needs. Data analysis involved collaboration between creative arts team members and social science team members who were highly experienced in qualitative data analysis. Our Young People’s Advisory Group were included in data interpretation.

We recognise that the positionalities of our research team shaped study delivery and data interpretation. Many team members identified as neurodivergent, some as non-binary, and almost all reported personal experience of at least one ACE. Some members were from rural places, often excluded in research, and some identified as being from an ethnic minority. These positionalities informed our sensitivity to issues of inclusion, emotional safety, power, voice and accessibility. Reflexivity was supported through individual reflective journals and team reflexive discussions consistent with established qualitative approaches (Sarfo & Attigah, 2025; Vicary et al., 2017). While the team was unified by a deep, shared commitment to youth voice, lived experience and the epistemic value of ABMs, we were alive to the challenges of working in interdisciplinary ways (we reflect upon these in Bhui et al., 2025). We created formal and informal spaces (e.g. workshops, analysis meetings and reflective conversations) to examine shared and divergent assumptions and values, and how we might resolve these. Our learning from these processes are documented in greater depth elsewhere in Batool et al., (2026). Our approach to researcher reflexivity aligns with our choice of data analysis method (Framework Analysis) and theoretical position (critical realist) (Mercier et al., 2023).

### Recruitment

Ethical approval was granted by the University of Oxford Medical Sciences Inter-Divisional Research Ethics Committee (R71941/RE001) and the NHS Health Research Authority Committee (23/WM/0105). Adolescents were eligible to take part if they: were between 10-24y; had experienced at least one ACE; were interested and able to contribute to a series of arts-based, group workshops exploring their experiences and mental health; and self-reported to be well enough to take part. In line with Alessi et al.’s (2023) trauma-informed research principles, we gave careful thought to building trusting relationship with gatekeeper organisations for recruitment, taking time for multiple meetings to fully explain the project, how young people had shaped it and to explore how we could best work with the organisation to learn from them about recruiting and supporting young people in research. We mostly worked with organisations and schools known to project team members, spanning the five English Attune study sites (Cornwall, Kent, London, Leeds, and Oxford) representing urban, rural and coastal regions. We especially sought underrepresented groups, including refugees, gypsy and traveller communities, LGBTQ+ groups and adolescents with learning disabilities. We took tailored our recruitment processes to individual young people, including multi-media materials and multiple meetings to enable them to get to know us and to decide if they wanted to take part.

All recruited participants provided informed consent, with additional parental consent for those under 16. Following consent, participants were invited to complete a survey which was administered to all young people taking part in the Attune project. This included a battery of measures primarily to understand their ACE histories and current mental health status of participants who had opted to take part in Attune; this is described in detail elsewhere (Bhui et al., 2026). The survey also asked participants about their preferences for art modalities in the creative workshops, and what the team could do to ensure they could take part safely, and with agency and enjoyment. Survey outcomes were only shared with the workshop facilitators if necessary (e.g. safeguarding, adjustment needs).

### Participants

Participants were N = 62 young people, with a mean age of 17.39 years (SD = 3.64), representing nine ethnic groups. They were recruited from nine diverse settings: N=9 from a higher education (HE) neurodivergent (ND) group; N= 16 from an LGBTQIA+ community group for diverse gender and sexual identities 16 participant; N= 6 from a refugee community group; N= 4 from a traveller and ROMA community group; N= 8 from a HE animation group; N= 3 from a coastal HE setting; N= 3 from a coastal college for adolescents with special educational needs (SEND); N= 4 from a young carers group and N= 9 from an inner city secondary school.

The complete demographics of all participants’ are reported in Supplementary Materials Tables S1 and S2. In summary, N = 44 identified as British and N = 30 as being from minoritised ethnic backgrounds. Of those that reported gender (N = 71), N = 33 (46.5%) identified as a woman/girl, N = 20 (28.2%) identified as a man/boy, N = 10 (14.1%) identified as non-binary, N = 19 (25.7%) identified as trans and N = 8 (11.3%) participants identified as ‘other’ genders (e.g., agender, autigender, gender fluid). A clinical diagnosis of a neurodivergence was reported by N = 23 (31.1%). The most commonly reported neurodivergent experiences (diagnosed and suspected but undiagnosed) included autism (N = 28, 37.8%) and ADHD (N = 24, 32.4%). N = 36 (48.5%) were in school or college. More than a third (N = 26 participants, 36.1%) reported two or more maltreatment experiences and 10% (N =8) reported four or more. The most common reported maltreatment was emotional abuse (N = 35, 47.5%), followed by emotional neglect (N = 25, 33.8%) and physical abuse (N = 20, 27.0%), respectively. N = 8 (10.9%) reported ever living in foster care Mean depression and anxiety scores were in the moderate severity range, with N =20 participants (27.2%) reported severe depression and 23 (31.08%) reporting severe anxiety.

### Designing Arts-Based Workshops

Establishing safety and trust in the research environment is a key principle of trauma-informed practice and spans physical, relational and internal safety (Alessi et al., 2023). A programme of workshops was developed, informed by our work on relational ethics (Pavarini et al., 2021), our analysis of the typical structure of community arts-based programs for young people (Williams et al.,2023) and Baim’s model of the Drama Spiral for working with personal stories (Baim, 2017). Priorities were safe, accessible, transparent and carefully paced work, tailored and responsive to individual young people, embedded with autonomy, choice and personal boundary-setting to live out our commitment to their individual choice and power. Everything was an invitation and tailoring, changing or opting out of activities was made easy. Workshops were designed to move from carefully scaffolded, externalised, low-risk and indirect engagement, through symbolic and representational levels, toward more personal recounting and meaning-making if appropriate, supporting internal safety (Alessi et al., 2023).

Consideration of the physical environment was integral to workshop design. Workshop sites were selected collaboratively with gatekeepers who could advise on suitable settings. Most workshops took place in schools and community venues routinely accessed by young people, while a small number were held in specialist settings, including a group therapy environment for particularly vulnerable participants and studio spaces on university campuses for older young adults and some refugee participants. Across all locations, quiet spaces were available. Participants’ preferences regarding physical and sensory environments were explored through the on-boarding Attune survey, and via an early creative den-building activity that examined safety, comfort and belonging, which informed subsequent physical workshop space design.

For the workshops, we assembled a diverse portfolio of expressive arts activities encompassing visual, performing, and media arts to maximise participant choice and accessibility. Participants could either engage in a single creative modality across their series workshops, choosing from animation, photography, or dance, or take part in structured, multimodal series of workshops incorporating collage, installation design (e.g., den building), mask making, animation, creative writing, drama, music, and film. Our trauma-informed practice included: a rigorous safeguarding policy, consultation with trauma specialists and access to support if needed, facilitator training, transparent and advance participant information on activities, imbued with choice, options and quiet zones, neurodiversity training and neuroinclusive environments with adaptations for communicating and/or sensory differences. Workshops were planned for groups of approximately ten participants at a time, who may or may not have known each other. Our aim was to support inclusion by accommodating a broad range of artistic orientations, skill levels, and communication needs linked to learning disabilities and language confidence (e.g. situational mutism). To ensure coherence across sites, all workshop programmes were underpinned by equivalent conceptual structures to ensure the creative practices practically and analytically aligned with our research questions set out in Table 1.

**Table 1.** Arts based workshop series in one project site showing media forms and example creative conversation prompts and interview questions.

| Week | Focus | Multi-modal Media | Creative Conversation Prompts | Interview Questions and links to Research Questions (adapted in context) |
| --- | --- | --- | --- | --- |
| 1 | Building trust, group relations and safety | Den making | What would your safe place be like? What would help you feel safe and comfortable in this group? | Where do you feel safe now, and where have you felt safe in the past? What is it about those places that helped you feel safe? (RQ3 & 5) |
| 2 | Creative tools for senses and memories | Sensory picnic (food art) | Are there colours you associate with your childhood and why? Can you think of a positive memory associated with this smell / taste/ texture? | What difficult life events can affect young people and how? What strategies do young people use to manage these experiences? What support do they need? (RQ 1, 3, 4 & 5) |
| 3 | Creative tools for storytelling | Exploring and making meaningful objects | What does this object mean to you? Tell us your story about this object | How would you explain your mental health to a friend, family member or professional? What has affected your mental health? (RQ3 & 4) |
| 4 | Planning Personal Stories | Collage (individual or group) | What images, colours or materials have you chosen to show important parts of your story? Which part of your collage feels most important to you and why? | What role has your childhood played in your 'journey' / growing up? What has been helpful and unhelpful to your wellbeing? (RQ2 & 4) |
| 5 | Composing Personal Stories | Masks and Shields | Do masks and shields have a place in real life? How might a mask or shield help or protect someone in real life? | What could protect young people, and their mental health? If you could create anything to support young people's mental health, what would you create? (RQ4 & 5) |
| 6 | Sharing Art as | Curating and | What would you like people to | Has using art helped you to express |
|  | Data | reflecting on created art | understand from your artwork? What do you hope people understand about your story through your artwork? | yourself, and in what ways? What is the most important thing you will take away from the workshops, and what helped you feel safe sharing with the group? (RQ6) |

### Workshop Delivery and Data Collection

Participants were invited to workshops in their area, with details of what was involved and who would be there. Each workshop lasted 2-3 hours and was facilitated by at least one arts practitioner and one researcher. Training to ensure standardisation across facilitators included developing a shared code of practice and protocol for data collection, regular meetings across sites to co-produce and compare workshop structures, and supervision by an identified research lead.

Participatory practice-based workshops (activities using creative media) were interspersed with two types of planned narrative data collection. First, participants were invited into ‘creative conversations’ connected to the creative activities, often as a shared small group discussion during art making. Facilitated by the art practitioner, these aimed to build on the opportunity provided by the creative activities to begin to explore associated participants’ experiences. Second, young people were invited to step out of the creative process for short, one-to-one interviews (5-10 minutes at a time) with a researcher. Interview questions were standardised across settings, but semi-structured in delivery, and explored the research question of focus in that week’s workshop. Table 1 shows a sample of ‘creative conversations’ and interview questions across the workshop series in one study site which used multi-modal media. All ‘creative conversation’ prompts and interview questions are reported in Supplemental Files. Support was offered to individual young people where needed through a safeguarding process aligned with local and national guidance. With permissions, all individual and group ‘creative conversations’ were audio-recorded for later transcription and analysis, and creative outputs were photographed and stored. Facilitators also generated field notes during the workshop and in debriefing session, focusing of the capturing workshop activities and context to situate the qualitative ‘creative conversations’ data, and to support researcher reflexivity. A worked example of workshop delivery in one region is provided in Supplemental Materials.

Workshop recordings of individual interviews and group discussions were transcribed verbatim, using pseudonyms created by participants. Other identifying details were removed. Artistic outputs were filed and tagged to the relevant timepoint in the transcript but were not subject to direct analysis. We have published elsewhere on the challenges and opportunities for analysis of artistic outputs by young people in mental health research (Shaughnessy et al., 2026).

### Framework Analysis

Data were analysed using Framework Analysis (FA, Gale et al., 2013) chosen for its suitability to catalogue and analyse large qualitative datasets via multiple coders as well as its flexibility for both inductive and deductive coding. We drew up Parkinson et al.’s (2016) worked example of applying FA to the study of mental health. A team of six analysts included workshop facilitators, other Attune researchers and purposively recruited coders. This interdisciplinary group worked collaboratively, starting in dyads with a workshop facilitator and someone who had not been in the workshop. The dyads and wider group met monthly to support consistent, transparent and auditable analytic processes. In stage 1 of FA (familiarisation), coders checked transcripts for accuracy and began deep reading, documenting initial observations in open coding to generate a close connection to the data. Stage 2 involved developing a thematic framework ready for deductive coding. To avoid confusion with other qualitative approaches, we use the term codebook to describe our thematic framework. The codebook was initially structured around our six a priori research questions, termed categories for the purpose of FA. Working independently followed by discussion in dyads, analysts conducted line-by-line coding, first assigning narrative segments to a best-fitting category and then inductively generating a descriptive code to capture its nuance. Under one or more of the six categories, new codes were added to a live codebook if they had been identified in at least two transcripts. Code meanings / definitions were added to the codebook with indicative quotes. Working with a live codebook meant all analysts were working with the most up-to-date codes. Weekly team meetings ensured cross-team agreement and use of codes, including merging codes. Codebook creation was an intensive process lasting approximately six months and generated 176 initial codes.

Stage 3 of FA (indexing) involved re-coding all transcripts according to the final codebook for consistency and completeness. In Stage 4, we charted the dataset by tabulating participants (rows) by categories and codes (columns), retaining links to supporting texts. This supported our within-category analysis and prepared the ground for collective interpretation work in Stage 5 (mapping and interpretation). Via collaborative and iterative group discussions and consensus, we generated final and evidential themes representing participants’ accounts of mechanisms linking childhood adversities to their current mental health and wellbeing.

Facilitators took these themes, supplemented with the artwork produced by our adolescent participants, back to participant groups my means of an in-person visit and a presentation. Participants were invited to respond (comment, question, refine) the data interpretation, and the facilitator made written notes of feedback. Revisiting was not possible with some participants no longer attending the settings (e.g. refugee group). Participants suggested only minor changes to the interpretation (e.g. questioning or resisting terminology such as resilience). What was evident from these sessions was the validation offered by seeing their creative works in relation to the project as a whole and realising the extent of the shared experiences across diverse young people.

### Methodological Integrity

We drew upon Guba & Lincoln’s (1994) criteria for evaluating quality. Credibility (whether the interpretations represent participants’ meanings and experiences well) was supported by multiple modes of sustained and deep engagement in data collection and analysis. In relation to data collection, our prolonged, trauma-informed time with young people supported trust that expressions of lived experience would be safe and validated. Our ABMs supported young people’s communication of lived experience in ways that were participant-driven, freely chosen and self-directed, and therefore likely to be authentic. ‘Creative conversations’ in workshops enabled deep inquiry to understand context and complexity in young people’s experience. These practices supported data credibility, in that young people expressed meaningful lived experience with minimal researcher influence. In relation to data analysis, credibility of interpretation was supported through extensive and systematic iterations of coding and continuous interpretative dialogue across coding teams to question, ground and evidence interpretations in the data (researcher triangulation). Whilst the creative outputs of the workshops (e.g. collages, dance, film) were not subjected to analysis in and of themselves, the data analysis teams continually connected to these to deepen familiarisation and enrich interpretation in ways that remained intimately connected with the creative context in which the data was produced. Reflexive practices helped continual awareness-raising of different researchers’ engagement with data interpretation, and emerging analyses were brought to the wider whole Attune team to interrogate the evidence for claims, which also supported confirmability. Participants’ own words are presented in the findings to ensure interpretations remain grounded in lived experience.

Transferability is supported through detailed description of the study context, participant characteristics, workshop structure, creative methods employed and analytic procedures, enabling readers to assess the relevance of the findings to similar populations and settings. Dependability was enhanced through the systematic application of FA, and the assigning of a ‘master controller’ who monitored all iterations. We documented the development of the analytic framework and coding definitions, staying close to participants’ language. Analytic decisions were recorded through meeting notes and directly onto the master codebook, providing a traceable account of how findings were generated. Together, these strategies ensured that the findings are grounded in the data while acknowledging the interpretive role of the research team.

## Results

### Overview

Our full analysis addressed six research questions and this paper reports findings for research question four: What are the mechanisms that young people believe are driving the way their ACEs come to affect their mental health? A central finding was that adolescents did not conceptualise mechanisms as simple, linear pathways linking ACEs to mental health outcomes. Their accounts focused more on how present-day experiences and contemporaneous conditions were saturated with determinants and mechanisms shaping their current mental health. We define determinants as factor (e.g. poverty) that influences the likelihood of an outcome (e.g. poor mental health). We define mechanisms as the processes or pathways (psychological, relational, or contextual) through which determinants exert effects. Findings adolescents’ own perspectives of what they felt were the mechanisms of these proximal, and not historical, determinants on their mental health and wellbeing. We use the conceptualisation of mental health and wellbeing of our participants, as emerged through the workshops: mental health and wellbeing is less about the absence of symptoms and more about feeling safe, understood and accepted; being able to express thoughts, feelings and needs; and having opportunities to develop a positive sense of self, feel you belong and have hope for the future. We begin with an overview of how our conceptual model of the findings, followed by participant accounts of the first two mechanisms.

Our conceptual model shows two broad categories of mechanisms: (1) System Failures and (2) Seeking Restoration (see Figure 1). System Failures represents intersecting interpersonal, environmental and relationally mediated intrapersonal mechanistic pathways causing harms to adolescents’ mental health. These harms were enacted through institutions and adult authority, particularly in schools and services meant to support them, and hence are understood as failures of the systems around the young person. In contrast, Seeking Restoration represents intersecting interpersonal, environmental, and relationally mediated mechanisms pathways that young people sought to recover from diverse harms, and involved pursuing experiences of embodied and relational safety to restore feelings of calm, self-worth and belonging. Interpersonal mechanisms refer to relationships with adults and peers, environmental mechanisms refer to material and sensory qualities of places and spaces and relationally mediated intrapersonal mechanisms refer to within-person psychological responses to experiences.

**Figure 1:**
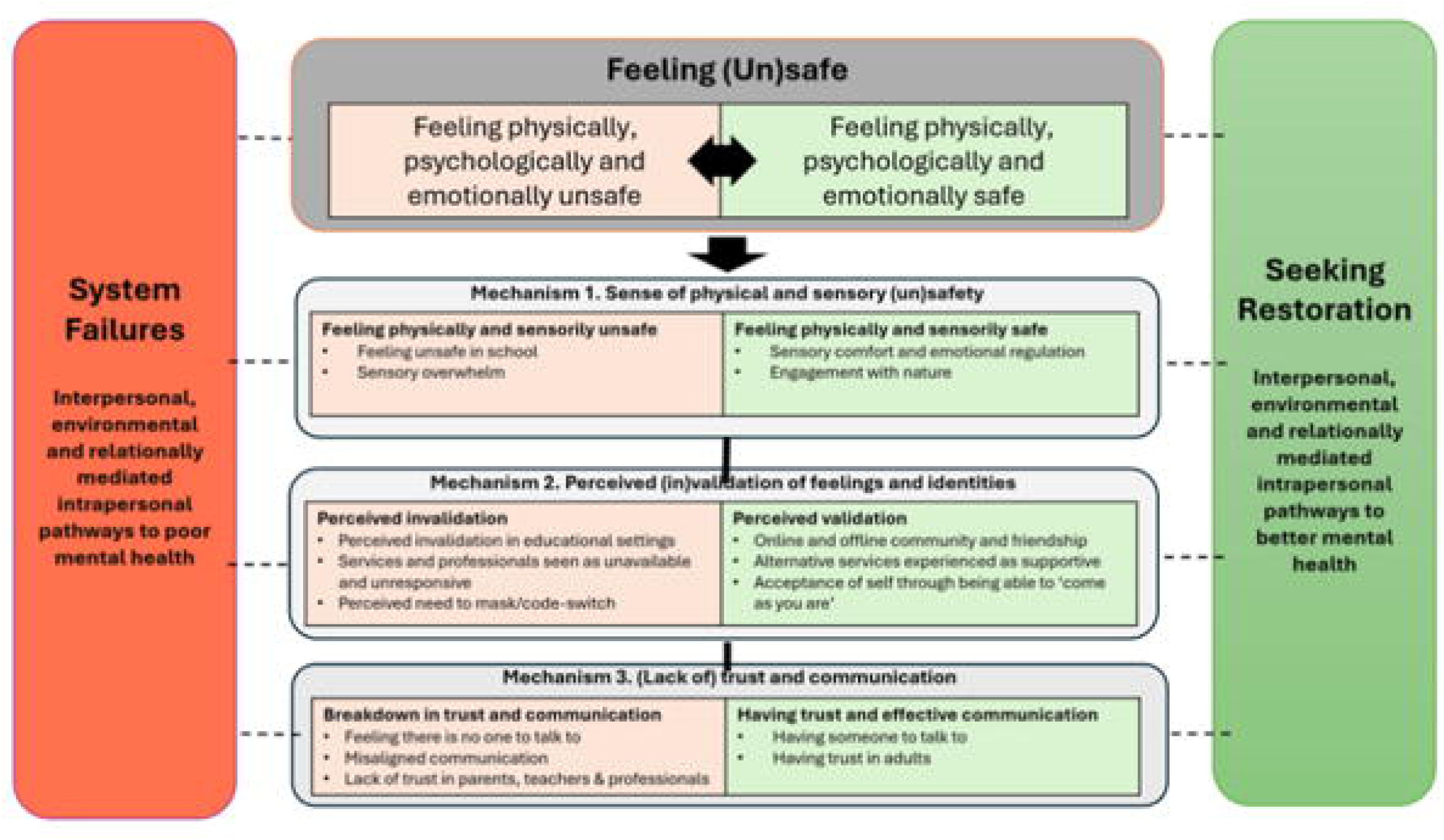
Adolescents’ accounts of mechanistic pathways influencing their mental health

The centrepin of the conceptual model is Feeling (Un) Safe. Across the dataset, adolescents’ accounts of how System Failures and Seeking Restoration influenced their mental health were consistently organised through the concept of safety, i.e., feeling safe or unsafe. Hence, Feeling (Un)Safe is the model’s central organising mechanism. By ‘feeling safe’ adolescents meant a place-based, embodied and relational experiences in which they felt calm, regulated, understood, accepted and protected. By ‘feeling unsafe’, participants meant the reality or expectation that they may be physically or emotionally harmed, rejected or dismissed though places and people. Adolescents conveyed three distinct aspects of their lives where Feelings of (Un)Safety worked mechanistically to affect their mental health, emerging either from System Failures or Seeking Restoration. These are categorised in the model as sub-mechanisms of (Un) Safety. Sub-mechanism 1 is ‘Sense of physical and sensory (un)safety’ and includes physical and sensory experiences, events or processes. Sub-mechanism 2 is ‘Perceived (in)validation of feelings and identities’ and includes legitimisation or disavowal of young people’s feelings and identities in educational and professional settings. Sub-mechanism 3 is ‘(Lack of) trust and communication’ and comprises experiences, events and processes relating to trust and communication. This is not presented here due to word limits. We present each the first two sub-mechanisms in turn, with illustrative quotes from the across the dataset. For each quote, we report the pseudonym chosen by the participant, their age range, and the group/ setting from which they were recruited. Identifiers for the quotes are as follows: ND HE = neurodivergent HE group; LGBTQIA+ group = community group for diverse gender and sexual identities; RF = refugee and asylum seekers’ group; Gypsy = traveller and ROMA community group (preferred term Gypsy), Coastal HE Animation Group = CHEA; CHE = Coastal HE setting; City School = inner city secondary school; SEND College+ = Coastal specialist college; NW = Central England young carers’ group.

### Sub-Mechanism 1: Sense of Physical and Sensory (Un)Safety

Physical and sensory experiences played a significant role in adolescents’ wellbeing. Participants’ accounts covered experiences of physical and sensory (un)safety emerging from System Failures, in in school (1.1) and via sensory overwhelm (1.2) and how, in response, they were Seeking Restoration though sensory experiences that restored comfort, regulation and wellbeing (1.3, 1.4).

#### 1.1 Feeling unsafe at school

Many participants reported that school was a place in which they felt unsafe: “a place that doesn’t make me feel safe is school”’ (Tilly, 18-25, Gypsy). Feeling physically and/or psychologically unsafe was mostly due to regular exposure to bullying, with some adolescents refusing to attend school as a result:

You can get bullied a lot over being, like, a Gypsy in high school…I don’t go to high school, because of that (Leah, 18-25, Gypsy).
You get stared at, like they wanna get you…[it makes me feel like I have] a lot of vulnerabilities (AH, 11-16, City School).

For BB, from the LGBTQIA+ group, school was a profoundly unsafe place:

BB: I have a lot of unsafe places… Mainly school…I hate my school.
Researcher: So what is it about the school that is, that feels, unsafe to you? Is it the people?
BB: Yeah, very much the people. Because one them wants me, wants me to kill myself

The sense of being unsafe and vulnerable was intensified for some young people by a feeling of no escape. Several participants drew on imagery of entrapment and imprisonment when describing school environments: “there is nowhere to run to in school, you have to stay” (AH, 11-16, City school). Georgia (18-25, Gypsy) described feeling ‘imprisoned’ in that setting that was harming her:

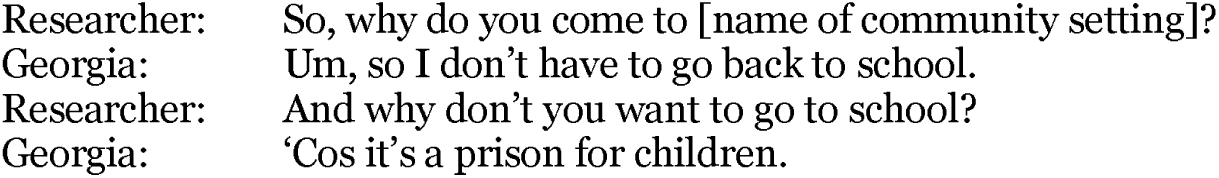

Bullying was also an experience of powerlessness and invalidation, especially when authority figures failed to keep them safe or respond to their distress (System Failures).

> get bullied a lot, through from primary school to secondary school. And I told teachers and they say, ‘oh, just stay away from them.’ But you can’t really stay away from them […] you either bump into them, or they come to you. So, they won’t do anything (Glitter, 11-15 LGBTQIA+).

Some participants attributed staff inaction to prioritising the school’s reputation over their safety whilst others reported that school staff blamed them for the victimisation:

> The ultimate aim [of headmaster] was protecting the school’s reputation… because when I went back to school the next day, people who were bullying me were still there. There’s no change” (Siproites, 18-25, ND HE).

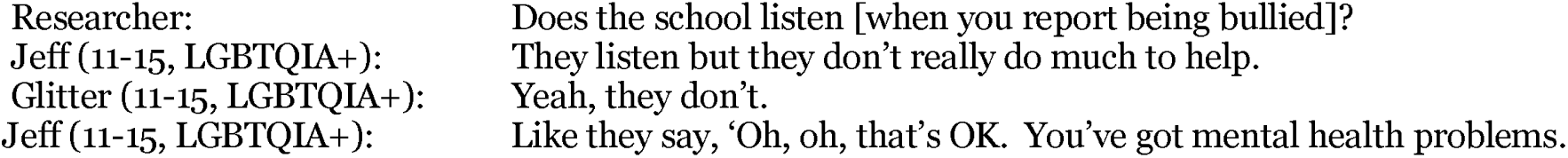

Adolescents’ perception was that schools did not recognise that bullying meant that school was a relentlessly unsafe place for them, with no route to safety, physically or via adult intervention. It was the combination of threat and failure to act that led young people to conclude that adults could not or would not protect them, in turn generating feelings of social abandonment. Young people also appeared to interpret institutional inaction as carrying symbolic meaning about their value; participants frequently concluded not only that they were unsafe, but that their suffering was insufficiently important to warrant protection from adults. Distress therefore also appeared linked to experiences of devaluation.

#### 1.2 Sensory overwhelm

Sensory overwhelm emerged across multiple sites and groups, indicating that feelings of unsafety were not restricted to interpersonal relationships but were also produced through embodied environmental experiences. Sensory overwhelm was often linked to loud noises, crowding and bright lights, as explained by the following young people:

> [I can’t] mentally switch off [at school] because there’s still going to be people around, there’s still going to be bright lights, white walls. There aren’t places where you can really mentally switch off” (Starfire, 18-25, NDHE).
>
> Ghost (16-25, LGBTQIA+, neurodivergent): Loud places…. like even that room, it’s really echoey and loud. And even small noises are really loud.
>
> Lee (16-25, LGBTQIA+): Rooms with lots of people.
>
> Vern (16-25, LGBTQIA+): Yeah, crowded places.
>
> Hindu (16-25 LGBTQIA+): No room to move.

UFO described fluorescent lights at school as physically and psychologically overwhelming, and the invalidation and ‘gaslighting’ of their experiences:

> a sensory trauma, I think it quite… there’s a lot of gaslighting and invalidation with sensory experiences. For example, so I’m very sensitive to light. Erm, but I didn’t know people experienced senses differently. So I used to be really confused why everyone else was okay with the pain from the lights. You know, everyone else seemed to be able to deal with it, but I couldn’t. And people used to be annoyed at me wanting the lights off, or hiding from the light. (UFO, 18-25, ND HE)

They also explained how they sorted to hiding under a blanket as a barrier against the pain from lights and they reported that were not going to school anymore as it is “very overwhelming”. Sensory overwhelm therefore functioned as a direct pathway into feelings of “not being able to deal with it” and subsequent unsafety. School was not the only institutional setting where artificial lighting caused significant sensory distress. Taran reported that the lighting in child and adolescent mental health services (CAMHS) inpatient settings impaired their ability to concentrate and benefit from therapy:

> I don’t think they think about, like, the room that you’re in. Not even like group sessions, but also the lighting. So for five years in CAMHS there was only one session where I had lights that weren’t fluorescent and flickering. And that session was one where we both came out of it being like oh, that was positive. The woman was like, oh I feel like we’ve made a real breakthrough. We hadn’t, but it felt… And I could remember what happened in the session. Because with the lights and the sensory things, I wouldn’t be able to remember what we talked about in sessions after they finished. So if we did come up with anything useful, I wouldn’t be able to put it in place. (Taran, 18-25, ND HE, and LGBTQIA+).

Like Taran and UFO, many participants did not distinguish sharply between physical, sensory and psychological safety. Noise, crowding and artificial lighting were experienced as assaults on their capacity to regulate attention, emotion and bodily comfort, and the lack of professional’s recognition of this was highly observed by young people, including Fred in his experience of the health and care system:

> the sensory stuff, especially in like, in the care system and like the NHS. Some of the ways that they would deal with like, non-autistic… They deal with all children the same, which can be a problem. Because some things that wouldn’t be traumatic to non-autistic children are very, very traumatic to autistic children, I n a sensory sort of way.” (FR, 18-25y, ND HE)

In these ways, sensory distress became particularly harmful when institutions failed to believe or accommodate it. The mechanism was therefore not sensory sensitivity alone but the interaction between sensory needs and environments perceived as inflexible or unresponsive. Thus, physical and sensory experiences were rarely described as just conditions but as communicating whether they were protected, recognised and able to exist safely.

#### 1.3 Sensory comfort and emotional regulation

In response to sensorily unsafe environments and being powerless to change them, adolescents described Seeking Restoration by exercising agency in shaping or withdrawing into preferred environments (Seeking Restoration). Minimising overwhelming sensory stimuli and the opportunity to withdraw from, or adjust, sensory input was foundational for many participants in fostering embodied safety. Comfort objects, preferred textures, smells and spaces functioned as self-generated forms of regulation that restored a sense of control and predictability. Comfort was described by participants as feeling good and at ease, physically and mentally, in the moment. Saron, an asylum seeker/refugee (16-24, RF), described her blanket as:

> Something [that] makes me [feel at] home […] It is really nice, it’s fluffy, it’s very comfort[ing]. So that one is mine, I don’t really offer it [to people].

Similarly, several autistic young people reported feeling comforted by the feel and/or smell of a soft toy, for example: “It’s more about the feel than the smell, so it has to feel right. I haven’t washed it in a long time” (Siproites, 18-25, ND HE); “The smell of that [teddy bear], like, holding it to my face and inhaling is, like, a comfort supercharge” (Six, 18-25, ND HE). For these participants, comfort objects were not merely sentimental keepsakes but active coping tools, contributing to emotional and psychological wellbeing in ways that were sensory specific and personally meaningful. Several participants also described the size and configuration of a space as influencing their sense of comfort and safety. For some, larger spaces were associated with ease and freedom of movement, “it just feels like […] if there is more space I would feel, like, more comfortable” (Saron, 16-24, RF), whilst for others, enclosed spaces provided a feeling of containment and security:

> In addition to engaging with I find, like, open spaces too overwhelming. Like, there’s too much happening, too much choice […] I like feeling […] enclosed, like a shoebox. I find it doesn’t feel constricting. It feels like a safety blanket. (Ducks, 18-25, ND HE).

As well as selecting sensory comfort objects and accessing open or enclosed spaces, young people described actively shaping or selecting environments to enhance comfort and reduce sensory overload, “the lights being off, and everything being quiet, and having the quiet room over there […] helped me a couple of times.” Georgia (18-25, Gypsy) emphasised that quiet, warmth, and lack of crowding were central to making a space feel safe and comfortable for her:

> A safe space has to be quiet, or else everybody gets chucked out the shed … it has to be warm […] It definitely has to be comfortable … and not a lot of people.

Several young people described using their school’s sensory room as a sanctuary to reduce stress and enable them to stay in school. For Fred, it was both the prevision of this space and teacher recognition of their need for it, that was helpful:

> Not all schools should have a sensory room because I don’t know if I’m just like the odd one out, but our school had a really good sensory room. And they […] would give you a card that you could leave and go to sensory room (Fred, 18-25, ND HE, in care).

However, not all schools provide these spaces, and UFO could no longer attend school due to sensory overwhelm. Since leaving school, they reported no longer needing to hide under a blanket, attributing this to being in a home environment that did not assault their senses:

> I don’t hide underneath it [blanket] so much anymore because … we don’t turn lights on now or we have natural light, not the proper big [fluorescent] light and stuff. So, I don’t feel there’s much need to hide under it. (UFO, 18-25, ND HE)

Across accounts, comfort objects and preferred environments were repeatedly characterised by personalised touches and predictability in their psychological affordances, underscoring how sensory control and predictability may be an important component of feeling safe. Psychological benefit appeared to arise as much from this restored sense of control as from the sensory properties themselves. Our participants’ accounts suggest that comfort was not simply a hedonic experience. Rather, feeling physically comfortable appeared to signal that they belonged within a given environment and that their experiences and needs were believed, and could exist without apology or challenge.

#### 1.4 Engagement with nature

Restoration from unsafety and distress was also actively sought by adolescents via time in nature. Whereas human systems were often characterised by judgement, demand and invalidation, nature was experienced as accepting, predictable and both psychological and sensorily safe. For some adolescents, positive sensory experiences intersected with appreciation for the natural environment, with participants stating that engaging with nature through direct sensory interaction, such as lying on the grass or feeling the rain, facilitated mindfulness and enhanced their ability to remain present and to feel safe, which they found beneficial to their mental health:

> Sometimes I just go lie on the grass in the rain and I literally just feel that moment … it completely changes your head state. It completely changes where you’re at. And I just think, I think it’s one of the things that is not used often enough in mental health (Yoda,18-25, Coastal HE).

This suggests safe sensory experiences (grass, rain) were a pathway into the benefit of nature which in turn contributed to a shift in mental perspective and overall psychological well-being:

> When I, I feel, um, sad or when I have something problems, when I go to nature, I feel better. Or when I go to mountain, I feel good (JU, 16-25, RF).
>
> When I felt like I was at the absolute lowest point possible … it didn’t matter what the weather was or what I had to do, I immediately put my jacket on and went out into the forest. Because it’s, kind of like, those trees, they will always be there … we’re all so caught up in our own world and our own problems that, when you about it, doesn’t actually matter (KA, 18-25, HEA).

Accounts of forests, mountains and rain also suggest that engagement with nature provided temporary relief from the monitoring, masking and vigilance that participants felt were required in their everyday social environments. Nature therefore appeared to create opportunities for psychological restoration through freedom from interpersonal threat (especially bullying), as well as via cognitive and emotional distancing from distress (‘switching off’).

Sub-mechanism 1 suggests that physical and sensory (un)safety was a present-day mechanism shaping mental health. Importantly, environments exerted influence not simply through objective features such as bullying, noise or lighting, but through the meanings attached to those experiences. System Failures were unsafe environments and unresponsive adults communicated a young person’s vulnerability, powerlessness and exclusion. Seeking Restoration was characterised by young people taking their own distress seriously and using their agency and intuition to source experiences to bring regulation and safety. Mental health was therefore conveyed as being dynamically shaped through relations and control between young people and the physical, sensory and interpersonal environments they inhabited.

### Sub-Mechanism 2: Perceived (in)validation of feelings and identities

A second (sub) mechanistic pathway shaping adolescents’ mental health centered around their experiences of perceived validation and invalidation. Participants’ accounts suggest that, to them, invalidation meant that their knowledge of themselves was challenged, discounted or overwritten by others. Across schools, services and everyday interactions (2.1, 2.2.), participants described experiences in which adults and institutions appeared not to trust their accounts of their own emotions, identities, needs or capabilities, driving them to mask (2.3). These were seen as System Failures whose psychological impact arose from having their legitimacy as knowers of their own experience undermined. In response, young people reported Seeking Restoration through more affirming relationships and communities (2.5, 2.6).

#### 2.1 Perceived invalidation in educational settings

Invalidation was a recurrent interpersonal mechanism described by participating young people. They especially characterised school as detrimental to their mental health largely due to perceived invalidation at the hands of peers and teachers, as explained by Tilly (18-25, Gypsy)

> Tilly: A place that doesn’t make me feel safe is school, ‘cos it’s just […] judgemental.
>
> Researcher: So, what about school feels judgemental?
>
> Tilly: The teachers. Teachers.

Morpheus and Starfire both reported having their expressions of feelings and anxieties invalidated and even punished by teachers at school:

> He [teacher] goes ‘I don’t want you using it [card signalling need to leave the classroom]. You’re too smart. This is a trend. What is this anxiety?’ I ended up feeling awful. I felt absolutely awful because I’d just had like a major panic attack and punishment for its expression (Morpheus, 18-25, ND HE).
>
> Often, I get quite emotional, like crying, and then you almost get into trouble for your reaction (Starfire, 18-25, ND HE).

In both accounts, emotional distress was redefined by teachers as a personal failure at best, and disruptive behaviour at worst. In these ways, adults were seen by the young people as self-appointed gatekeepers of emotional legitimacy, effectively determining whether their distress was warranted, acceptable, or deserving of adjustments in school. This is extended in the case of Sonic (18-25, SEND college), who recounted multiple ways in which his schools privileged their own reading of him above his own understanding of his autistic identity, needs and abilities:

> The school told my parents ‘this boy will never get his GCSEs, he’ll never get far in life, he won’t do anything.’ That’s quite upsetting because they’ve given up on you. When I was younger, I was in a corner for my first, up until year 5 […] because the teachers didn’t know what to do with me … pushing you in the corner […] playing with toys instead of actually giving the lessons I needed.
>
> What they [teachers] find easy, we [neurodivergent young people] find very difficult

Other participants also reported that their needs and concerns were dismissed by teachers, including learning plans being ignored, personal coping strategies disallowed and sensory needs denied:

> The amount of times I’ve brought up that it [Individual Learning Plan] exists, the teachers have looked at me like I’ve grown a third head (Ducks, 18-25, HE ND).
>
> [I’m] not allowed to wear headphones [to block overwhelming noise experienced in school corridors], only if you have a diagnosis, but I wish I was (Potato, 12-18, LGBTQIA+).

Yoda was frustrated with the ways that schools placed significant pressure and stress on students, and yet when they expressed that stress, they responded in emotionally invalidating ways, orienting to the young person as being dramatic or losing perspective:

> I used to go into school and worry about a piece of homework. I’d go in, have a complete breakdown, and the teacher would be like, ‘it’s not that important’ (Yoda, 18-25, CHE).

Across account, invalidating experiences were experienced by some young people multiple times per day at schools, and hence are positioned as System Failures influencing wellbeing. Invalidation challenged young people’s confidence in their own emotions, identities, needs and interpretations of events, often leading to self-concealment, distrust and feeling unsafe to be themselves. When expressions of anxiety, panic or sadness are met with invalidation, criticism and even punishment, it can contribute to masking (2.3).

#### 2.2 Services and professionals as unavailable and unresponsive

Invalidating encounters with professionals and services were also described by young people as negatively shaping their mental. Participants often described services as reproducing rather than alleviating feelings of powerlessness as professionals’ interpretations were privileged above their lived experience and was particularly acute within services whose stated purpose was care. In these ways, they were seen as System Failures. Several participants described support services as insufficiently youth-centred or responsive. The participant called Six, for example, described CAMHS as prioritising professional agendas over listening to him, leaving him feeling unheard and dismissed:

> CAMHS…It was just a system that was very much not adapted to hearing what you had to say but centred on what they gathered from what you said. So, they could ignore what they wanted to and fill in what they wanted to and do whatever they felt was best…there’s just a lot of professionals who would deny anything I said (Six, 18-25, ND HE).

Six’s account suggests that distress was amplified not only by unmet need but by experiences of epistemic disqualification. Participants described professionals as occupying a position of interpretive authority in which young people’s own accounts of their feelings, needs and experiences could be questioned, revised or replaced. Mental health was therefore shaped not solely by problems requiring support, but by feeling unable to influence how those problems were understood. Many participants from marginalised groups also reported that services were ill-equipped to understand their specific experiences. For example, UFO described how no recognition of their neurodivergence complicated their access to effective support, and in effect re-traumatised them:

> So, I’ve experienced a lot … because of not knowing I was neurodivergent … also because of like an incident that had something to do with that as well. But those services add so much trauma on to it. Which is ironic in a way [laughs] (UFO, 18-25, ND HE).

Another young person described seeking professional help in a crisis and being turned away, leaving them feeling that their distress was not urgent or serious enough to warrant immediate support:

> I was on the phone to the crisis line…you know, the, the very end people that you need to talk to, and they left me on hold for three hours. And so, it’s like, you’re at that point in your life where you need someone desperately… um, and if it wasn’t for my mum, you know, it wouldn’t have been a good outcome, because they left me for three hours on hold when I was in a crisis. (Yoda, 18-25, CHE)

Yoda’s interpretation was that service responsiveness communicated the legitimacy and urgency of his distress, and deservingness of support. Taken together, across 2.1 and 2.2, participants did not describe settings or services as harmful simply because support was unavailable or ineffective. Rather, harm arose when young people felt that those settings communicated that young people’s experiences, identities or needs were not fully recognised, understood or taken seriously.

#### 2.3 Perceived need to mask

Many participants described feeling compelled to mask or in environments they perceived as unsafe, judgemental, or discriminatory. Masking refers to the conscious or unconscious suppression, modification, or concealment of aspects of oneself to avoid negative judgement, rejection, discrimination, or social sanction (Miller et al., 2021). Participants described masking as altering behaviours, emotions, interests, communication styles, or neurodivergent traits to appear more acceptable to reduce the risk of harm. Across accounts, masking appeared less a matter of personal preference than adaptive responses to anticipated invalidation. Participants often modified their self-presentation because they expected expression of their real self to expose them to judgement, misunderstanding or rejection. In this way, experiences of invalidation operated prospectively rather than retrospectively. Young people did not need to be actively criticised in a particular moment; prior experiences of non-acceptance were sufficient to shape future behaviour, encouraging self-monitoring, concealment, and emotional labour.

Several participants described adapting how they presented themselves in school and other social settings to remain safe. For example, Star Light described the emotional labour of increasing their masking following an autism diagnosis:

> … after the [autism] diagnosis, I think it became more of, like, a survival mood or kind of a trauma response, because it [masking] was kind of, like, okay, now this is the reason why I’m finding x, y and z so difficult. So, now I’m gonna, like, mask to protect that side of me. And I think that’s where, like, it then takes a bit more of an emotional toll (Star Light, 18-25, ND HE).

For Star Light, greater awareness of difference heightened their concerns about how others might respond. Masking functioned as a protective strategy. This suggests that experiences of invalidation may not be only directly shaping wellbeing, but also via the sustained effort required to anticipate and manage this. For example, KE (18-25, CHE) described concealing his gender identity at school because of fears about how others would respond:

> it was very tough […] being in, like, the school environment and feeling very, like, hidden. Like obviously [I] couldn’t present myself […] I was presenting myself as male, but I couldn’t like literally say, ‘oh hey I’m a guy’ to teachers or anything. So obviously everyone was calling me the wrong name and the wrong pronouns, and yeah, I was not doing well (KE, 18-25, CHE)

KE’s account highlights how schools can communicate powerful assumptions about identity through everyday interactions and institutional norms. The distress described here did not arise solely from overt hostility, but from the continual experience of being disavowed personal choices. The persistent gap between authentic identity and socially recognised identity appeared psychologically burdensome, contributing to feeling “very hidden” with diminished wellbeing. Other participants reflected on the longer-term consequences of sustained masking. Groot (18-25, ND HE), for example, described the profound tension involved in attempting simultaneously to embrace and conceal his autistic identity. He explained that he was “trying to start a new chapter as my authentic [autistic] self, whilst also masking to be like everyone else”. Reflecting on the impact of this tension, he noted: “Either way, I would get isolated. I got lost within the mask.”

Thus, experiences of anticipated invalidation encouraged young people to monitor, alter, and conceal aspects of themselves to navigate environments perceived as unsafe. Rather than functioning simply as a social strategy, masking also appeared capable of disrupting young people’s relationships with themselves. Participants described how prolonged concealment could erode authenticity and create uncertainty about who they were beneath the performance required by others. Distress therefore appeared linked not only to social exclusion but to the psychological costs of maintaining a separation between public and authentic selves.

#### 2.4 Alternative sources experiences as supportive

In contrast to System Failures which invalidated young people, our participants reported actively seeking their own restorative relational experiences. Central to these was feeling accepted and being able to be themselves. First, some young people identified alternative settings and services as more responsive and culturally sensitive than mainstream educational or clinical institutions. Participants from the refugee cohort, for example, highlighted the role of charities in providing meaningful support:

> Like, [name of charity] is an important charity […] lots of young people are going to it for help, for anything, because they have a lot of trouble with social workers or Pas [Personal Advisors, housing specialists for 18+ care leavers]. So, the importance is [name of charity] is working hard to help. It’s, it’s, it’s a good charity (Mr. Lucky, 16-24, RF).

Alternative therapies were also described by some adolescents as being beneficial:

> With the lady that I went to see privately, she did art and music therapy alongside, kind of, normal talking therapy, and I found that that was massively helpful to me (Yoda, 18-25, ND HE).

#### 2.5 Online and offline community and friendship

A second context in which participants described Seeking Restoration was through friendships and community. Whereas schools and services were often described as settings in which experiences, identities, or needs were questioned, community appeared to support wellbeing through a mechanism of acceptance and belonging. Participants described finding people who shared identities, experiences, interests, or values and that these reduced feelings of difference and young people did not need to justify or defend who they were. Their restorative value therefore appeared to derive less from the absence of adversity and more from the presence of recognition and acceptance, which afforded needed experiences of safety. For example, many adolescents described the importance of “finding your tribe” in schools, youth groups, neighbourhoods, and online spaces. One participant explained: “all the people I like the most are there [youth club] and I feel safe there” (Ant, 16-18, LGBTQAI+).

Community settings were frequently described as places where participants could feel part of something, reassuring them that they were not “alone in your own experience” (Siproites 18-25, ND HE). For some young people, particularly those from marginalised groups, this sense of belonging appeared especially important because it countered experiences of exclusion. For some refugee participants, community groups appeared to serve functions ordinarily associated with family and kinship networks. In contexts marked by displacement, separation, and uncertainty, community organisations were experienced as providing continuity, care, belonging, and social anchoring, as explained by Ju:

> It’s nice to have staff [in the community group] and other young people as well. We build that relationship. I feel like I belong to them, and we feel like we are a family because we don’t have family here” (JU, 18-25, RF).

Similarly, Saron contrasted her experiences in a community with that of clinical therapy:

> For me to improve my mental health I don’t feel like therapy works for me personally. Community does work. I feel like I want to belong [with] someone. I like to have someone who is like-minded, who can understand me […] who can listen to me […] I have done therapy before, but I don’t feel they understand me most of the time” (Saron, 18-25, RF).

Notably, participants did not reject professional support per se. Rather, their accounts suggested that community was valued because it offered reciprocity, mutual understanding, and shared experience - not afforded in clinical approaches. Furthermore, unlike many formal helping relationships, which position one person as helper and another as recipient of help, community relationships were characterised by mutual recognition and solidarity. It was this experience of being understood “from the inside” that appeared particularly restorative.

Digital spaces extended opportunities for belonging beyond geographical and social boundaries. Participants described fandoms, gaming communities, social media, and online networks as spaces where they could connect with others who shared interests, identities, or life experiences, which in turn helped them to feel safe:

> Through anime and video games I found community where I can be safe […]” (Sonic, 18-25, SEND college).
>
> I feel like, if anything, social media can be really helpful because people can talk about it […] the more people talk about how they cope with it, the more other people start to understand how they might be able to cope with it” (Leia, 12-18, City School).

Digital communities were more than sources of entertainment. For some young people, online spaces broadened access to validation and support that was difficult to find in their immediate offline worlds.

A notable feature of these accounts was that trust appeared to emerge from belonging rather than precede it. Once young people felt accepted within a friendship group or community, they became more willing to disclose vulnerabilities, seek support, and express emotions openly. Trust therefore seemed to function as a mechanism through which belonging translated into wellbeing benefits, as Muzan and Morpheus explained:

> I just feel comfortable with people here, that’s why I’m always expressive…” (Muzan, 18+, SEND College).
>
> Sometimes it’s not like a benefit [being autistic], but it teaches you the people who do deserve to be in your life. So, like this group, for example, where we can be open and honest with each other and we can wear whatever we want, do whatever we want (Morpheus, 18-25, ND HE).

Feeling recognised by others appeared to reduce the need for masking and self-protection. Some participants also described supportive digital friendships and vicarious relationships, such as connections with online creators, fictional characters, or public figures who openly discussed mental health and adversity. Seeing others disclose struggles, share coping strategies, and normalise distress helped participants feel less alone. Validation from peers appeared to contribute to stronger self-worth and more positive self-concepts, laying the groundwork for the deeper processes of self-acceptance described next.

#### 2.6 Acceptance of self through being able to ‘come as you are’

As a third type of Seeking Restoration from invalidation, many participants described seeking greater self-acceptance. Rather than emerging solely through individual reflection, self-acceptance appeared to develop through participation in relationships and communities where young people felt recognised, understood, and able to express themselves without fear of judgement. In this way, self-acceptance functioned as a relationally mediated intrapersonal mechanism through which supportive communities influenced wellbeing. Several participants reflected on the importance of being understood whilst simultaneously recognising the limits of mutual understanding. Eivor described how connection could arise through shared experience without requiring complete sameness:

> Someone that understands the pain, the suffering, everything […] Me and Sonic have gone through similar experiences, but they will never be exact. Like we can never truly understand what each other feels or have gone through (Eivor, 18-25, SEND College)

Eivor’s reflection suggests young people value being understood enough. Self-acceptance was not achieved through conformity to a group identity but through finding spaces where individuality could safely coexist with belonging. For some participants, broader experiences of social acceptance appeared closely linked to self-worth and self-acceptance. Max, for example, connected hopes for citizenship and belonging within wider society to feeling accepted as a person:

> I’m here and I want to make my life here. I want to live here and like become a citizen, hopefully. So that’s definitely going to help me feel accepted for who I am, yeah (Max, 18-25, CHEA).

Max explained that his being accepted by wider communities was inextricably linked to his own self-acceptance. Identity exploration around sexuality and gender was described by several young people as other important aspect of this process of self-acceptance. Participants spoke about moving from uncertainty, concealment, or confusion towards greater authenticity, often supported by affirming relationships and communities. Minnie reflected:

> I came out as trans when I was quite young. I was like, 13, but… to my family and close friends. But only recently I started, like, my public transition, and I’ve been telling people at school and at school nights, and I just feel so much better. I can be myself, so, it’s like, don’t hide (Minnie, 12-18, LGBTQIA+).

The improvement in wellbeing described by Minnie appeared linked not simply to disclosing an identity but to no longer having to conceal it. Consistent with participants’ earlier accounts of masking, psychological distress appeared to arise when aspects of self had to be hidden or managed. Conversely, being able to inhabit and express an authentic identity appeared intrinsically restorative. Self-acceptance therefore functioned partly through reducing the psychological burden associated with concealment and self-monitoring. For others, participation in community settings enabled the reclamation of previously hidden aspects of identity and selfhood. UFO described how feeling they belonged in a supportive community allowed them to reconnect with interests that had previously been masked:

> In the community I am able to reclaim interests that I used to mask and not want to share with people […] I feel like I have grown a lot since becoming part of the community (UFO, 18-25, ND HE,).

Notably, UFO frames growth not as acquiring something new but as recovering parts of themselves that had previously been suppressed. This suggests that self-acceptance may involve a process of reclaiming identities, interests, and ways of being that were abandoned or concealed in response to anticipated invalidation. Community settings appeared to facilitate this process by creating environments in which authentic self-expression was experienced as safe rather than risky. Acceptance from others appeared to scaffold acceptance of self.

Across 2.3, 2.4 and 2.6, the young people’s accounts suggest that self-acceptance functioned as a key restorative mechanism linking belonging and validation to improved mental health. Supportive communities appeared to enable young people to move beyond merely feeling safe towards feeling able to exist without the need for hiding, changing or apologising, and by having opportunities to express and be oneself.

## Discussion

This study aimed to generate new, lived-experience understandings of the mechanistic pathways through which ACEs shape adolescent mental health and wellbeing, as defined by them. Drawing on our large qualitative dataset generated through ABMs with 62 highly diverse young people in England, the findings offer an understanding of mechanisms from young people’s perspectives. A key finding is that, rather than focusing on how historical ACEs and trauma affect them now, adolescents foregrounded the dominant influence of present-day relational, environmental, and contextual conditions shaping their mental health. We first consider how these findings may be understood in terms of dominant ACE frameworks and reflect on why participants may have decentered ACEs as influential in their current mental health. We then discuss young people’s accounts, as represented in our conceptual model, before we offer implications for practice and research.

### Comparison with ACE frameworks

Much of the ACEs literature has focused on cumulative exposure, dose-response relationships, and associations with later outcomes (e.g., Hughes et al., 2017; Jia & Lubetkin, 2024). That work has been transformational in establishing the long-term significance of adversity and associations with mental health outcomes. Whilst undisputed, our findings suggest that adolescents may conceptualise mental health through a different explanatory logic to that which dominates much ACE research. They rarely organised their accounts around discrete past adverse events or circumstances, nominating mechanisms linked to what is happening to them now rather than what happened in the past.

We consider four possible reasons why participants did not produce narratives of historical trauma-to-outcome as expected. First, it may have been that our methods were not sufficiently safe or supportive of recounting ACEs and tracing their mechanistic unfolding over time. Second, the immediacy of participants’ present circumstances are likely to have shaped which experiences were foregrounded during the workshops (although many did recount experiences from childhood). Third, while ACEs may have influenced their present mental health, their effects may have yet to be accessed with sufficient narrative coherence to emerge in our approach. We return to the implications of these methodological considerations below.

Finally, however, the decentering of ACEs does align with a developmental psychopathology (DP) framework (Cicchetti & Toth, 2013). DP conceptualises mental health outcomes as emerging from biological, psychological, relational, and environmental influences across development rather than from isolated adverse events operating through simple causal pathways. Within this perspective, ACEs are understood as important developmental experiences that can shape trajectories, but their effects are neither fixed nor deterministic (Hawes & Allen, 2023). This view helps explain why participants in the present study rarely described past adversity as having a direct bearing on their lives now. At the same time, and without contradiction, it is also likely that these present-day experiences were inflected by the legacy of ACEs. A DP perspective would posit that the salience of present-day safety could be understood as a proximal process through which earlier adversity continues to exert influence, although we must remember that this was not how our young people conceptualised it. It is, in many ways, to be expected that orientations to (un)safety would be high among those whose past adversities have engendered feelings of vulnerability. Indeed, this is the premise of trauma-informed practices - to acknowledge the likelihood that trauma and adversity infiltrate a person’s self-concept, ways of being in the world and what they need now (Yatchmenoff et al., 2017). The implication of the DP framing is that our findings may indeed represent mechanisms linking ACEs to adolescent mental health, but that these are operationalised by sustained exposure to harmful environments and a sustained need for safety. A complexity here is that many of our participants were neurodivergent and their experiences of unsafety related to being in neurotypical worlds may layer on, or be intensified by, ACEs.

Bearing all of these possibilities in mind, we remain close to, and led by, the narratives of our young participants. We do not discount their narratives as artefacts of potential methodological limitations, nor as robust evidence of a DP explanation, but interpret them as reflecting their knowledge about the mechanisms shaping their mental health. We do this whilst remaining open to the potential continuing presence of ACE-related influences in these young people’s lives.

### Organising Mechanism: (Un)safety

Across the dataset, adolescents’ accounts of wellbeing were consistently organised through experiences of felt (un)safety. Despite substantial differences in identity, adversity exposure and presenting difficulties, participants converged in their emphasis on the extent to which they felt recognised and protected within their current worlds, hence its central position in our conceptual model. Safety encompassed emotional, psychological, sensory and identity-based dimensions. We therefore argue that felt safety may represent a transdiagnostic mechanism underlying adolescent mental health following adversity. Our conceptual model identifies two broad categories shaping felt (un)safety: System Failures and Seeking Restoration.

### Mechanism Category: System Failures

Schools, services and adult institutions frequently appeared not as contexts in which the effects of adversity were managed, but as contexts in which vulnerability was reproduced. Rather than being experienced as spaces of support and development, schools, mental health services, and other systems of adult authority often left young people feeling unheard, disbelieved, misgendered, pathologised, overexposed, or punished for expressing distress. For some neurodivergent participants, the sensory environment of school, including noise, crowding, and overstimulation, was described as profoundly dysregulating, particularly when requests for reasonable adjustments were dismissed. For others, experiences of bullying, stigma, breaches of confidentiality, or institutional inaction conveyed that their wellbeing was secondary to organisational priorities. Across diverse groups, participants consistently linked their mental health difficulties to environments in which they felt unable to be themselves, unable to trust adults, or compelled to mask aspects of their identity.

While some of these experiences, such as bullying (Man et al., 2022) and minority stress (Jardas et al., 2023), are well documented as harmful to young people, our findings extend this literature in several important ways. First, they highlight the central and under-recognised role of invalidation as a mechanism of harm. Participants repeatedly described experiences in which their feelings, identities, or needs were dismissed, minimised, or reframed by adults. This can be seen as a form of epistemic injustice, in which young people’s knowledge of their own experiences is discounted. Invalidation not only undermined their self-understanding but conveyed they were unworthy of care, intensifying their distress. Although emerging research has begun to examine invalidation within services and interpersonal contexts (e.g., Cunningham et al., 2024; Wasson Simpson et al., 2022), it remains largely absent as a central construct in youth mental health research. Our findings suggest that invalidation is not only pervasive but also, in principle, preventable, making it a critical target for intervention in youth mental health work.

Second, our findings contribute to the emerging literature on sensory overwhelm, particularly in educational environments, as a significant but often overlooked source of distress for neurodivergent young people (e.g., Pavlopoulou et al., 2025). Participants described sensory experiences not simply as uncomfortable, but as highly dysregulating and, at times, unsafe. They conveyed how their mental health was deeply embodied in the environmental, with noise, lighting, and crowding functioning as chronic stressors. They centered sensory experience as a core causal mechanism shaping their current mental health and are being more widely documented in the literature (Price & Romuladez, 2025; Taels et al., 2023), contributing to calls for more intervention in young people’s sensory experiences (Yuan et al., 2022).

Third, while the relationship between masking and mental health in neurodivergent populations is increasingly recognised (e.g., Chapman et al., 2022; Ross et al., 2023), our findings extend this by situating masking within a broader ecology of unsafety. Masking may be better understood as an indicator of environmental unsafety than as an individual behavioural characteristic. Participants rarely described masking as a preferred strategy. Instead, masking emerged as a rational adaptation to contexts perceived as threatening, invalidating or punitive. This shifts attention from asking why young people mask to asking what kinds of environments make masking necessary. From this perspective, masking becomes evidence of relational and institutional failure rather than evidence of individual vulnerability. In sum, masking was an emotionally costly response to avoid harm. As it constrained young people’s authenticity, in conditions of overwhelm or fear, the need to mask could be another key mechanism linking harmful systems to adolescent mental health.

The centrality of (un)safety within participants’ accounts is notable, theoretically coherent and clinically relevant. Feeling unsafe, whether physically, emotionally, or psychologically, is an inherently destabilising experience with clear implications for mental health (Lynch et al., 2025). Indeed, Maslow defined safety as a ‘metaneed’ (Maslow, 1954, p. 8). When individuals do not feel safe, they may experience heightened anxiety, increased vigilance to threat, and a reduced capacity for emotional regulation, connection and creativity (Brosschot et al., 2018). In this sense, unsafety constrains core processes associated with psychological wellbeing, including openness to experience, authenticity, relational connection, and growth (Kashdan & Rottenberg, 2010). Crucially, our findings suggest that the impact of unsafety for young people is intense when it is experienced within contexts that are expected to provide care, protection, or support, including schools and youth services – a finding also being reported by others (e.g. Mori et al., 2021; Horanicova et al., 2024). As noted earlier, it is plausible that participants’ emphasis on present-day unsafety reflects, in part, an understandable sensitisation to threat shaped by earlier experiences. Even so, our findings extend beyond this by highlighting that ongoing, contemporary experiences of unsafety are, to young people, mechanistically central to their mental health, rather than merely downstream effects of earlier adversity. Much work is now responding to the needs for settings, including schools and services, to be more accountable about their how adverse use of ‘power over’ young people is shaping their wellbeing (e.g. Berring et al., 2024; Corres-Merdano et al., 2025).

### Mechanism Category: Seeking Restoration

Against these harmful relationships and environments, young people described how they were Seeking Restoration towards safety and wellbeing. Their accounts challenge portrayals of adversity-affected adolescents as passive recipients of risk or resilience factors. Instead, participants acted as agents in their own wellbeing, intentionally seeking environments, relationships, sensory experiences, objects and communities that enhanced feelings of safety and belonging.

Restoration could be found in friendship, community belonging, online affinity spaces, supportive adults, contact with animals or nature, creative expression, and sensory regulation. These were actively pursued resources, reflecting young people’s agency and motivation to shape their environments, and their mental health, where possible. Also notable was the meanings attached to these. Participants described these experiences as places where they could feel safe, understood, less alone, and more authentic. Being in nature, as reported in other studies on youth wellbeing (Puhakka & Hakoköngäs, 2024), afforded a multisensory access to cognitive, emotional and embodied regulation. Belonging also emerged as particularly important for our participants, a finding being increasingly documented (e.g. Allen et al, 2024). Being with others who shared identities, interests, or experiences appeared to reduce shame and isolation while supporting self-acceptance and reducing the need for masking. This was particularly important for some of our marginalised participants (i.e. refugees, non-binary, neurodivergent). Being recognised and affirmed by others enabled more positive self-concepts and a stronger sense of worth. Again, these findings underscore how feeling safe was fundamental to the young people’s wellbeing, and that, this happens relationally rather than solely intrapsychically. This finding does not diminish the value of psychological therapies for ACE-affected young people (Moyes et al., 2022) but reminds us that restoration and wellbeing is also sought out by young people in ordinary relational contexts which afford companionship, recognition, acceptance and solidarity.

### Our Art-Based Methods

We noted earlier some methodological influences which may have limited the breadth of experiences, especially those directly about ACEs, which could emerge via our ABMs. Here, we reflect on the importance of our choice of ABMs for the population and subject matter. Our methods appeared to enable forms of expression and reflection that may have been less accessible through conventional interviews or checklist measures alone. Participants initially communicated through design, improvisation, images, movement, metaphor, music, objects, poetry, humour, sensory description, engaging individual and collaborative arts-making activities, and seemed to give the young people space and pace to talk about the meaning of these in relation to our research questions. Choice of these methods also reduced the chances of inadvertently reproduce adult-led perspectives. The positive reception of these methods by many participants suggests that they were not merely alternative data collection techniques, but part of the conditions that made participation possible.

The methods may also help explain the prominence of present-focused accounts. Creative practice is often grounded in process, immediacy, sensory attention, collaboration, and making something in the moment. Workshops may therefore have functioned as temporary spaces in which participants could step outside dominant trauma narratives centred exclusively on past harm and instead articulate what currently supports or undermines them. In doing so, they enabled accounts of identity, aspiration, resistance, and everyday coping that can be overshadowed in trauma research. For qualitative psychology, this indicates that methods can shape the quality and content of the data. ABMs may be particularly valuable where researchers wish to understand embodied, affective, and socially situated dimensions of psychological life.

### Implications

Our findings have implications for services working with young people who have lived through ACEs. First, trauma-informed approaches may benefit from complementing questions about historical adversity with questions about present-day safety. Assessments focused only on historical trauma risk overlooking, and perhaps legitimising, contemporary determinants of distress that may be modifiable now. Second, schools and youth-serving institutions should be recognised as potential sites of either harm or repair. Participants’ accounts suggest that relatively ordinary practices such as listening, respecting confidentiality, responding flexibly to distress, recognising identity, addressing bullying, and making sensory adjustments, may have substantial mental health significance. Conversely, dismissive or rigid responses may compound existing vulnerability. Third, interventions should take seriously the restorative resources identified by young people themselves. Community groups, peer spaces, creative opportunities, green spaces, and identity-affirming environments may not be peripheral extras but central components of mental health support. These resources may be especially important for marginalised young people whose experiences are poorly mirrored in mainstream settings.

Importantly, young people’s narratives suggest that developmental trajectories remain highly malleable during adolescence. Their descriptions of seeking belonging, sensory comfort, community, authenticity, and trust indicate active engagement with developmental resources that may counterbalance adversity and support positive adaptation. Consequently, our findings reinforce the value of moving beyond static ACE scores towards a developmental understanding that attends to the ongoing interplay between past adversity and present-day social ecologies in shaping adolescent mental health.

### Strengths, Limitations and Future Research

A strength of this study was the inclusion of a diverse sample of young people, many from groups underrepresented in adversity research, and the use of arts-based, participatory methods designed to enhance accessibility and agency. The large qualitative dataset and interdisciplinary analytic process enabled examination of shared patterns alongside individual nuance. Several limitations should be noted. Participants self-selected into a creative project and may differ from young people who would not engage in such settings. The data were cross-sectional and reflect accounts at one point in time rather than changing trajectories. Our findings concern participants’ meaning-making and should not be interpreted as showing that past adversity is unimportant. In addition, while creative outputs informed context and interpretation, they were not analysed as standalone data objects. Future research should continue to elicit young people’s perspectives on the causal pathways between ACEs and mental health, and how lived experience knowledge can and should be integrated with dominant psychomedical approaches to the study of ACEs and youth mental health. Research must reach and engage with diverse young people (e.g. based on ethnicity, gender and sexual identity, neurodivergence, and socioeconomic or educational position) to ensure that the evidence base informing policy and practice reflects the realities of more diverse young lives (Perowne et al., 2024).

## Conclusion

Young people in this study did not speak about mental health solely through the lens of past adversity. They more often described wellbeing as shaped by ongoing experiences of recognition, safety, belonging, authenticity, and the quality of their encounters with contemporary systems and relationships. These findings suggest that understanding adversity requires attention not only to what has happened in the past, but to what continues to happen now in their everyday lives. Interventions may therefore benefit from asking not only what adversities young people have experienced but also whether the environments they currently inhabit allow them to feel safe enough to flourish.

## Data Availability

Due to the sensitive nature of the data and ethical commitments to participant confidentiality, the qualitative datasets generated during this study are not publicly available. Anonymised excerpts supporting the conclusions of this article are included within the article and supplementary materials. Further information about the data is available from the corresponding author upon reasonable request and subject to ethical review.

## Author Contributions

SHJ contributed to the conception of the study, analysis and manuscript writing. CF contributed to study design, data collection, analysis, and manuscript writing. LA contributed to creative methodology development, workshop facilitation, data collection, analysis and manuscript preparation. AF contributed to creative methodology development, workshop facilitation, analysis and manuscript writing. EW contributed to data analysis and manuscript writing. KB contributed to study conceptualisation and manuscript writing. NS contributed to the conception of the study, creative methodology development, workshop facilitation, data collection, analysis and manuscript writing.

## Acknowledgements

We thank the young people who participated in this research and shared their experiences with us. We acknowledge the UKRI funding (MR/W002183/1) that supported this work through the MRC/AHRC/ESRC Adolescence, Mental Health and the Developing Mind Programme. We thank the numerous community organisations that facilitated recruitment and data collection, and the ATTUNE Young People’s Advisory Board for their guidance throughout the project. We acknowledge all ATTUNE team members who contributed to data collection and analysis across multiple sites and disciplines.

