## Supplementary Materials for "Mechanisms linking Adverse Childhood Experiences to adolescent mental health: a participatory arts-based study of adolescents’ accounts"

**Supplemental Material**

**Table S1**. *Sociodemographic Characteristics of Participants (n = 74)*

| **Variable** | **Value** |
| --- | --- |
| **Age (years), *M(SD)*** | 17.39 (3.64) |
| **Gender, n (%)** |  |
| Man/Boy | 20 (27.0) |
| Woman/Girl | 33 (44.6) |
| Non-Binary | 10 (13.5) |
| Other | 8 (10.8) |
| *Agender* | 1 (1.4) |
| *Autigender* | 1 (1.4) |
| *Genderfluid* | 4 (5.4) |
| *Genderqueer* | 1 (1.4) |
| *He/They* | 1 (1.4) |
| Don’t know/Not sure | 2 (2.7) |
| Prefer not to say | 1 (1.4) |
| **Trans Status, n (%)** |  |
| Yes | 19 (25.7) |
| No | 45 (60.8) |
| Don’t know/Not Sure/Questioning | 7 (9.5) |
| Prefer not to say | 2 (2.7) |
| **Ethnicity, n (%)** |  |
| African | 6 (8.1) |
| Arab | 3 (4.1) |
| Caribbean | 1 (1.4) |
| Chinese | 1 (1.4) |
| English, Welsh, Scottish, Northern Irish, British | 44 (59.5) |
| Gypsy or Irish Traveller | 2 (2.7) |
| Indian | 1 (1.4) |
| Pakistani | 2 (2.7) |
| Mixed/Multiple Ethnic Background | 6 (8.1) |
| *Mixed, White and Black African* | 1 (1.4) |
| *Mixed, White and Asian* | 3 (4.1) |
| *Mixed, Any Other* | 2 (2.7) |
| Other | 8 (10.9) |
| *Other, Asian* | 1 (1.4) |
| *Other, Black, African, or Caribbean* | 1 (1.4) |
| *Other, White* | 6 (8.1) |
| **Sexuality, n (%)** |  |
| Asexual | 5 (6.8) |
| Bisexual or pansexual | 22 (29.7) |
| Gay/Lesbian | 9 (12.2) |
| Heterosexual/straight | 19 (25.7) |
| Don’t know/not sure | 6 (8.1) |
| Prefer not to say | 3 (4.1) |
| Other | 10 (13.5) |
| **Neurodivergence, n (%)** |  |
| ADHD/ADD or Attention Differences | 24 (32.4) |
| Autism or Autism Spectrum Condition | 28 (37.8) |
| Demand Avoidance | 4 (5.4) |
| Dyscalculia | 6 (8.1) |
| Dyslexia | 14 (18.9) |
| Dyspraxia | 6 (8.1) |
| Intellectual/Learning Disability | 9 (12.2) |
| Mutism | 10 (13.5) |
| Language Impairment | 2 (2.7) |
| Synaesthesia | 5 (6.8) |
| Tourette’s Syndrome | 3 (4.1) |
| Not in education | 9 (12.2) |
| **Employment Status, n (%)** |  |
| Full-Time | 1 (1.4) |
| Part-Time | 14 (18.9) |
| Zero-hour contract/holiday job | 4 (5.4) |
| Not employed | 55 (74.3) |
| **Hunger Status, n (%)**  Not at all  Sometimes/Once or Twice  **Parents/Carers born in the UK, n (%)**  Yes, one parent  Yes, both  No  Unsure | 60 (81.1)  14 (18.9)    16 (21.6)  37 (50.0)  20 (27.0)  1 (1.4) |
| **Parent/Caregiver Status, n (%)** |  |
| Yes, more than one parent/caregiver | 51 (68.9) |
| Yes, one parent/caregiver | 16 (21.6) |
| No parent/caregiver | 7 (9.5) |
| **Foster/Residential Care Status, n (%)** |  |
| Yes, currently | 1 (1.4) |
| Yes, in the past | 7 (9.5) |
| No | 62 (83.8) |
| Don’t know/Prefer not to say | 4 (5.5) |
| **Parent/Carer Education Status, n (%)**  Not sure/Other  Attended secondary school  Completed GCSEs  AS/A2 Level  Technical/Vocational Training  Bachelor’s Degree  Master’s Degree  Advanced Degree/PhD | 23 (35.9)  4 (6.3)  6 (9.4)  2 (3.1)  3 (4.7)  20 (31.3)  4 (6.3)  2 (3.1) |

**Table S2**. Childhood Maltreatment and Mental Health Characteristics of Participants (n = 74)

| **Clinical Variable** | **Value** |
| --- | --- |
| **Childhood Maltreatment (SCMQ), M(SD)**  **Type of Maltreatment, n (%)**  Physical Neglect  Emotional Neglect  Physical Abuse  Emotional Abuse  Parental Violence  Sexual Abuse  Sexual Assault    **Number of Maltreatment Exposures (/7), n (%)**  None/Zero  1 exposure  2 exposures  3 exposures  4+ exposures    **Impact of Events (PTSD Symptoms), M(SD)**  Intrusion Symptom Severity  Avoidance Symptom Severity | **1.64 (1.91)**    10 (13.5)  25 (33.8)  20 (27.0)  35 (47.3)  8 (10.8)  10 (13.5)  10 (13.5)    24 (32.4)  9 (12.2)  9 (12.2)  9 (12.2)  8 (10.9)  **23.14 (12.96)**  11.20 (7.10)  12.08 (6.59) |
| **Formal Clinical Diagnosis Received** | 23 (31.1) |
| **Number of Neurodiversity Diagnoses, n (%)** |  |
| One diagnosis | 8 (10.8) |
| 2 diagnoses | 8 (10.8) |
| 3+ diagnoses | 7 (9.5) |
| **Depression Severity (PHQ-9/PHQC), M(SD)** | **13.25 (8.07)** |
| PHQ-9 = 0-4; No/Minimal Depression, n (%) | 13 (17.6) |
| PHQ-9 = 5-9; Mild Depression, n (%) | 10 (13.6) |
| PHQ-9 = 10-14; Moderate Depression, n (%) | 18 (24.4) |
| PHQ-9 = 15-19; Moderate-Severe Depression, n (%) | 12 (16.3) |
| PHQ-9 = 20-27; Severe Depression, n (%) | 20 (27.2) |
| **Anxiety Severity (GAD-7), M(SD)** | **10.93 (6.60)** |
| GAD-7 = 0-4; No Anxiety, n (%) | 13 (17.7) |
| GAD-7 = 5-9; Mild Anxiety, n (%) | 18 (24.5) |
| GAD-7 = 10-14; Moderate Anxiety, n (%) | 18 (24.5) |
| GAD-7 = 15-21; Severe Anxiety, n (%) | 23 (31.3) |
| **Mental Wellbeing (SWEMWBS), M(SD)** | **20.84 (6.12)** |
| **Loneliness (UCLA-4), M(SD)**  UCLA-4 = 4-6; No/Low loneliness, n (%)  UCLA-4 = 7-9; Moderate Loneliness, n (%)  UCLA-4 = 10-12; Moderate-High Loneliness, n (%)  UCLA-4 = 13-16; Severe Loneliness, n (%) | **10.68 (3.99)**  15 (20.3)  14 (19.0)  14 (19.0)  29 (39.3) |
| **Difficulties with Emotion Regulation Score (DERS), M(SD)** | **54.36**    **(17.14)** |
| DERS Awareness, M(SD) | 8.82 (3.44) |
| DERS Clarity, M(SD) | 8.51 (3.89) |
| DERS Goals, M(SD) | 9.22 (3.52) |
| DERS Impulsivity, M(SD) | 8.91 (3.74) |
| DERS Non-Acceptance, M(SD) | 8.95 (3.64) |
| DERS Strategies, M(SD) | 9.11 (3.75) |

**S3**. ***Examples of Workshop Researcher Interview Questions***

*Tailored to setting, artforms, group, age and context.*

**Workshop One**

- Where do you feel safe? Where have you felt safe in the past?
- What helps you feel safe?
- Are certain places that feel safe? What is your favourite place what is your least favourite place?
- Where would you like to live in the future? Why?
- What are the qualities that you most value in a place?

**Workshop Two**

- What difficult life events can we think of? [“that’s a good example of an ACE”]
- Have you or someone you know experienced one of these events?
- How did this/these affect you? How did you deal, cope, or manage this? Positive or negative?
- What do young people (like yourself) need to stop difficult life experiences having a negative effect on mental health? What would you say to professionals? Teachers? Family?

**Workshop Three**

- What do we mean when we talk about someone’s ‘mental health’?
- What does mental health mean to you? How would you explain your mental health to someone? A friend? A family member? A therapist/counsellor?

**Workshop Four**

- What role has your childhood played in your 'journey' / growing up?
- What has been helpful and unhelpful to your wellbeing?

**Workshop Five**

- If you could make anything to help young people and their mental health, what would you make – money no option.
- What do young people (like yourself) need to stop difficult life experiences having a negative effect on mental health? What would you say to professionals? Teachers? Family?

**Workshop Six**

- What is the most important thing you take away from this series of workshops?
- Has creative practice made it easier or more difficult for you to express yourself? What were the moments you felt most/least true to yourself? What helped/made it difficult for you to feel safe to share?
- What motivated and de-motivated you to share your artwork with others vs. keep it private?
- How have your relationships with others in the group changed throughout the workshops?
- What helped/made it harder to create feelings of trust throughout the experience?

**S4.** *Example of Workshop Delivery*

We provide detail of how flexible workshop delivery and data collection was operationalised at one regional study site in England, which included workshop delivery across five different settings. These were settings for refugees, young people with learning disabilities, an LGBTQ+ community group, autistic higher education students and a diverse multicultural school with a significant proportion of young people from deprived backgrounds. In Workshop 1 (Safe Spaces), participants were invited to create personalised physical spaces (dens) according to their sensory and creative preferences and which helped them to feel safe. ‘Creative conversations’ explored how environments (place, contexts), identity and socio-cultural positionings can be risky or protective of mental health and wellbeing, and what young people do to feel safe and mitigate harm. Workshop 2 (Sensory Picnic) offered food-based art activities to explore sensory experiences and food related memories and embodiment. ‘Creative conversations’ explored the concept of memories, and historical and personal narratives, and opened reflection on how young people define and experience adversity through embodied knowledge. For Workshop 3 (Objects) participants brought or described meaningful objects. ‘Creative conversations’ focused on tools (material or non-material) for emotional safety and resilience, why these are needed and their influence on mental health and wellbeing. Collage was the art form for Workshop 4. Here, participants narratively created and documented their journeys as well as future aspirations. ‘Creative conversations’ explored how young people define and explain mental health in relation to their histories and lived experiences. Workshop 5 (Masks / Shields) invited participants to create masks or shields using various materials. ‘Creative conversations’ explored themes of identity, protection, and authenticity, and where greater protection during and after ACEs could have helped them. Finally, Workshop 6 (Curation/Sharing Museum) was a collaborative reflection and future-oriented thinking sessions where participants curated their creative work for sharing and aimed to draw together collective understandings of histories of adversities unfold to shape their mental health.
